# A patient-centered view of symptoms, functional impact, and priorities in post-COVID-19 syndrome: Cross-sectional results from the Québec Action Post-COVID cohort

**DOI:** 10.1101/2023.05.27.23290638

**Authors:** Nancy Mayo, Marie-Josée Brouillette, Emilia Liana Falcone, Lesley K Fellows

**Author notes:** **Declaration of competing interests:** The authors have no completing interest related to this paper.

## Abstract

**Background:** Health services planning and mechanism-focused research would benefit from a clearer picture of symptoms, impact, and personal priorities in post-COVID-19 syndrome (PCS). This study aimed to provide estimates of the symptom, function, and quality of life (QOL) impact of PCS.

**Methods:** People living in Quebec, aged ≥18, were eligible for the Québec Action for/pour le Post-COVID (QAPC) study if they had symptoms lasting more than 4 weeks post-acute SARS-CoV-2 infection, with or without a positive COVID-19 test. Recruitment was through conventional and social media between September 2022-January 2023. Standardized and individualized questionnaires, in French or English, were accessed through an online portal. We report cross-sectional results from the baseline visit of the first 414 participants in this ongoing longitudinal study.

**Results:** Individuals spontaneously reported symptoms attributable to an average of 4.5 organ systems. Fatigue was most frequent. Effects on function and quality of life were moderate to severe, and had already persisted for a year or more in the majority. Personal intervention priorities included fatigue and post-exercise malaise, cognitive symptoms, shortness of breath, and impaired taste and smell. Women and men were similar on PCS impact, while older age was associated with lower impact.

**Interpretation:** Symptom clusters defined a range of severity, with fatigue a pervasive symptom at all levels of severity. Participants in this study are likely to be representative of those seeking health care for post-COVID-19 symptoms in Canada and the results can inform next steps for clinical, research, and health services planning.

## INTRODUCTION

As of January 2023, it is estimated that 4.5 million Canadians have been infected with SARS-CoV-2, including 1.3 million in Quebec.(1, 2) It became evident early in the pandemic that symptoms could persist or arise after the acute infection. In Quebec, the Institut National d ’Excellence en Santé et Services Sociaux recognizes post-COVID-19 syndrome (PCS) when symptoms last more than 12 weeks(3), the same time frame as the WHO(4). Prevalence estimates from population-based studies around the world range from 3 to 70% (5-14) depending on the study sample, timeframe, and methodological rigour. Statistics Canada estimates prevalence at 14.8%, some 1.4 million people(14). Symptoms also vary in nature and frequency across studies, with over 200 symptoms reported.(15) Few studies have reported on function, health-related quality of life (HRQL) or quality of life (QOL), but available evidence suggests substantial negative impacts. A recent structured review(16) found four studies reporting on the impact of PCS on quality-adjusted life-years based on the EQ-5D, finding a health reduction equivalent to losing 3 to 4 years of a hypothetical 10-year lifespan, values that are in the same range as chronic stroke, multiple sclerosis, or diabetes(17-19).

Facing a new and poorly understood health condition, the Quebec Action for Post-COVID (QAPC) study aimed to contribute a patient-centered understanding of symptom patterns, impact, and intervention priorities in a self-identified Quebec sample. The objectives of this initial report were to estimate the prevalence and severity of the health effects and life impact of PCS and the extent to which these differed by age and sex. In addition, while multiple symptoms can potentially affect those with the condition, we aimed to identify which of these symptoms are most bothersome for those with the condition, information that could guide the development of services to address these areas of priorities. The study used well-validated patient-reported outcome measures (PROMs), including the Patient Generated Index (PGI), an individualized measure suited to eliciting the most frequent and most bothersome symptoms)(20). This fully virtual bilingual study further aimed to empower participants by providing information regarding their own health profiles and access to self-management resources.

## METHODS

A cross-sectional analysis of the first 414 participants in a longitudinal study recruited from September 23, 2022 to January 31, 2023 was carried out involving people from Quebec who self-identified as having symptoms of the post-COVID-19 syndrome. The sample was assembled from multiple sources: most participants were reached through French- and English-language media (radio) and social media, with some contacted via email outreach to a waitlist for a post-COVID research clinic. Residents of Quebec age 18 and over were eligible if they currently had symptoms occurring 4 or more weeks post onset of symptoms of the COVID-19 infection, with or without a positive test.

### Procedures

The project (2022-8066) was approved by the Research Ethics Board of the McGill University Health Centre. People interested in participating were directed to the QAPC website to register. The study coordinator recorded their contact information, generated a study identification number and invited them into the study. Upon invitation, they were directed to an online web-portal “Research Electronic Data Capture” (REDCap) to enter their unique identification number, allowing them entry into the data capture platform. Following an e-consent process, they recorded their health outcomes information. All were asked to consent to open data sharing for secondary analyses and to be re-contacted for additional studies.

Recruited participants also were provided a password to access online self-management resources addressing breathing difficulties, cognitive symptoms, fatigue, and increasing physical activity. To manage mental health symptoms, participants were directed to BounceBack®, a free program offered by the Canadian Mental Health Association to help manage low mood, mild to moderate depression, anxiety, stress or worry(21).

### Measurement Framework

The Wilson-Cleary model guided the health outcomes assessment(22). A lightweight yet comprehensive approach used patient-centered assessment tools to characterize symptoms and their functional and quality of life impact at recruitment and over time.

As little was known at this study’s outset about the health effects beyond a symptom inventory(23), we adopted an individualized approach to outcome measurement. The Patient Generated Index (PGI)(20) asks people to nominate areas of their life affected by a health situation, here the sequelae of COVID. Each area is then rated on severity, from 0, “not at all”, to 10, “worst imaginable severity”. The person is then asked to consider areas where they most desire improvement and distribute a theoretical 10 tokens across the nominated areas. A total score is generated by multiplying the severity rating by the number of priority tokens allocated and summed. This process avoids the over-reporting that can occur when people are asked to choose from a list of symptoms, rather than spontaneously declaring them(24), and provides information about priorities in the person’s own words.

The only measure that has been recommended to date for assessing the health impact of post-COVID is the SF-36(25). We used it in its publicly available form (RAND-36)(26). We also used an internationally recognized health utility measure, the EQ-5D-5L(27, 28), which queries 5 functions: walking, usual activities, self-care, pain/discomfort, and anxiety/depression. These were supplemented with a series of visual analogue scales to cover the additional areas of fatigue, sleep, distress, shortness of breath (SOB), health rating, and overall quality of life (29, 30). Two additional single questions asked about fatigue impact (31) and minutes of exercise over a week. Questionnaires on post-exertional malaise (PEM) (32), post-traumatic stress disorder (PTSD)(33) and cognitive concerns(34) were also administered. Demographic information, co-morbidity, and COVID experience and vaccine history were also collected. Assessment tools had strong evidence for validity with robust measurement properties, were available in English and French, and in most cases had existing Canadian or Quebec norms(35-37) or comparative values(34)aiding interpretation. Each participant’s information was summarized, interpreted, and shared with them in the form of a personal dashboard, similar to one we had previously designed and tested for people living with HIV(38).

### Data Analysis

Descriptive statistics were calculated for each variable after recruitment of 100, 200, 300 and 414 participants and values on study measures did not vary across these waves suggesting that early and late responders did not differ on severity. Two people had missing data on language and on hospitalization. Where relevant, the effect of sex and age were estimated using linear, quantile (median), ordinal, or logistic regression depending on the distribution of the variable under study. When there was a sex effect, data were presented separately for men and women. For a sex effect to be considered important, the confidence interval around the estimated effect had to exclude the null value and the difference in prevalence between men and women had to be greater than 10%(39). Text threads were analysed using natural language processing (i.e., stemming and lemmatization).

Cluster analysis (k-means) was used to identify the extent to which the functional consequences of fatigue, cognition, sleep, distress, physical function, and shortness of breath clustered in individuals. For this analysis, all variables were converted to a score out of 100, with 100 being the worst level. The effects of the symptom burden of PCS were estimated using linear regression by regressing health and quality of life outcomes on cluster membership, adjusted for age and sex. Quantitative analyses were conducted using SAS V9.4.

## Results

**Table 1** presents personal and COVID-related characteristics of the sample of mean age 48.7 years, and predominantly women (75.8%). The hospitalization rate of 6% was similar to the rate in the Canadian population. Older people were more likely to be hospitalized that younger people and men spent more days in hospital than women. The vaccination rate of the sample was greater than the Quebec population. The date of last COVID event was reported for all but 3 of the participants and the average time from last event was 320 days. Reported substance use ranged from ≈5% for smoking and cannabis use to 26.6% for alcohol consumption and 16.9% reported increased consumption. More than ¾ of the sample were working prior to COVID and over 90% of those reported changing their working hours with PCS; almost half reported being on sick leave. Even with PCS, the sample reported carrying on with usual roles and responsibilities, with women having more household responsibilities than men. More than half had money challenges.

**Table 1.**
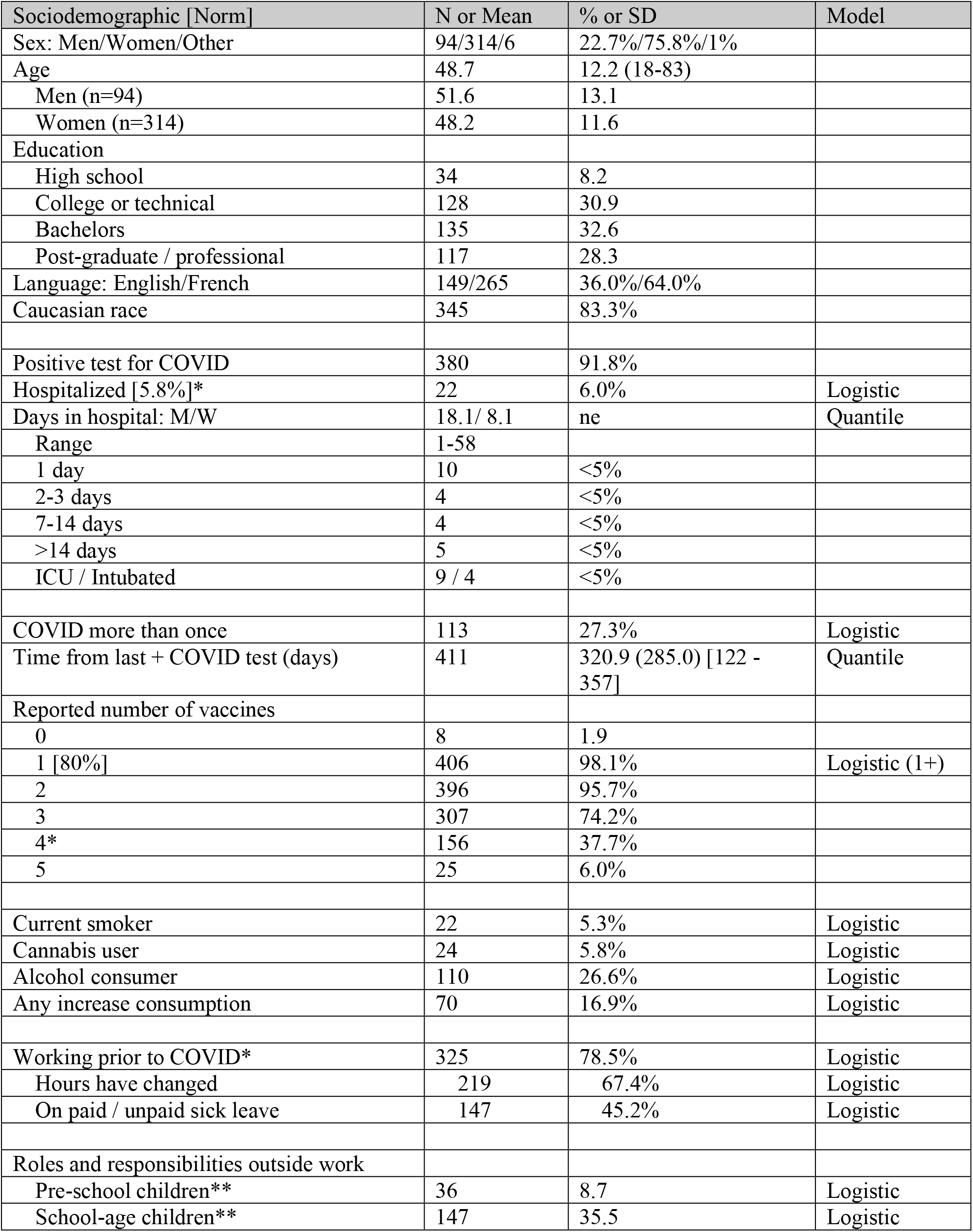
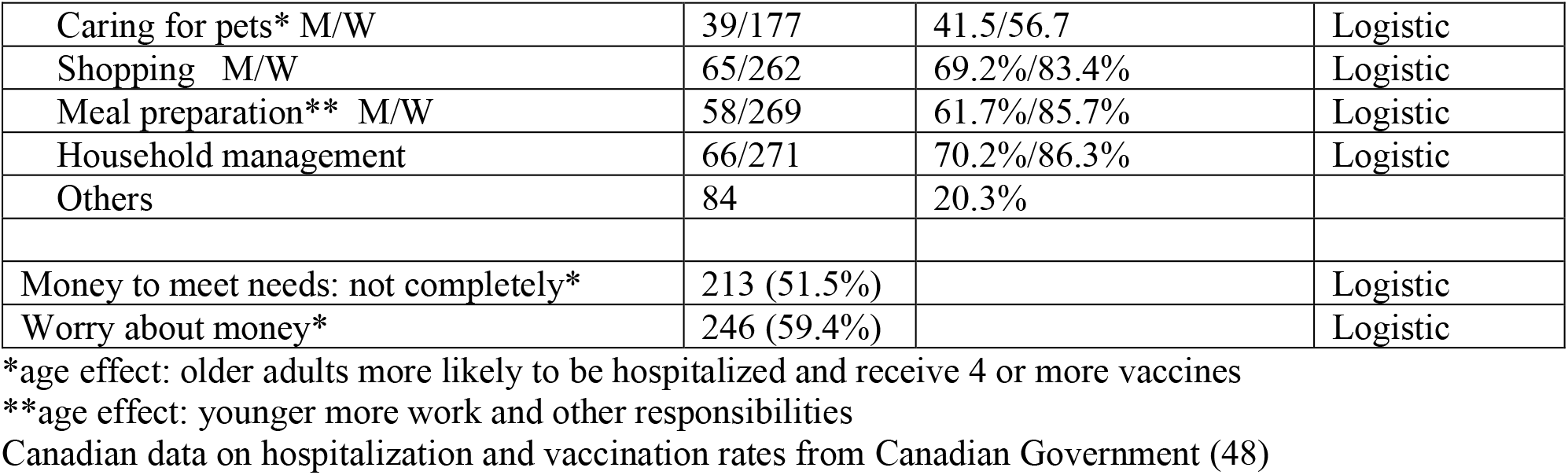
Characteristics of the Sample (n=414)

| Sociodemographic [Norm] | N or Mean | % or SD | Model |
| --- | --- | --- | --- |
| Sex: Men/Women/Other | 94/314/6 | 22.7%/75.8%/1% |  |
| Age | 48.7 | 12.2 (18-83) |  |
| Men (n=94) | 51.6 | 13.1 |  |
| Women (n=314) | 48.2 | 11.6 |  |
| Education |  |  |  |
| High school | 34 | 8.2 |  |
| College or technical | 128 | 30.9 |  |
| Bachelors | 135 | 32.6 |  |
| Post-graduate / professional | 117 | 28.3 |  |
| Language: English/French | 149/265 | 36.0%/64.0% |  |
| Caucasian race | 345 | 83.3% |  |
| Positive test for COVID | 380 | 91.8% |  |
| Hospitalized [5.8%]* | 22 | 6.0% | Logistic |
| Days in hospital: M/W | 18.1/ 8.1 | ne | Quantile |
| Range | 1-58 |  |  |
| 1 day | 10 | <5% |  |
| 2-3 days | 4 | <5% |  |
| 7-14 days | 4 | <5% |  |
| >14 days | 5 | <5% |  |
| ICU / Intubated | 9 / 4 | <5% |  |
| COVID more than once | 113 | 27.3% | Logistic |
| Time from last + COVID test (days) | 411 | 320.9 (285.0) [122 - 357] | Quantile |
| Reported number of vaccines |  |  |  |
| 0 | 8 | 1.9 |  |
| 1 [80%] | 406 | 98.1% | Logistic (1+) |
| 2 | 396 | 95.7% |  |
| 3 | 307 | 74.2% |  |
| 4* | 156 | 37.7% |  |
| 5 | 25 | 6.0% |  |
| Current smoker | 22 | 5.3% | Logistic |
| Cannabis user | 24 | 5.8% | Logistic |
| Alcohol consumer | 110 | 26.6% | Logistic |
| Any increase consumption | 70 | 16.9% | Logistic |
| Working prior to COVID* | 325 | 78.5% | Logistic |
| Hours have changed | 219 | 67.4% | Logistic |
| On paid / unpaid sick leave | 147 | 45.2% | Logistic |
| Roles and responsibilities outside work |  |  |  |
| Pre-school children** | 36 | 8.7 | Logistic |
| School-age children** | 147 | 35.5 | Logistic |
| Caring for pets* M/W | 39/177 | 41.5/56.7 | Logistic |
| Shopping M/W | 65/262 | 69.2%/83.4% | Logistic |
| Meal preparation** M/W | 58/269 | 61.7%/85.7% | Logistic |
| Household management | 66/271 | 70.2%/86.3% | Logistic |
| Others | 84 | 20.3% |  |
| Money to meet needs: not completely* | 213 (51.5%) |  | Logistic |
| Worry about money* | 246 (59.4%) |  | Logistic |

\*age effect: older adults more likely to be hospitalized and receive 4 or more vaccines
\*\*age effect: younger more work and other responsibilities
Canadian data on hospitalization and vaccination rates from Canadian Government (48)

**Figure 1** shows the number of areas nominated by people with PCS according to organ system. General systemic symptoms (which includes fatigue) were the most frequently nominated followed by symptoms related to cognition, pain, and the respiratory, cardiovascular and neurological systems. The symptoms with the highest priority for improvement were fatigue, cognition, respiration, taste, and smell.

**Figure 1.**
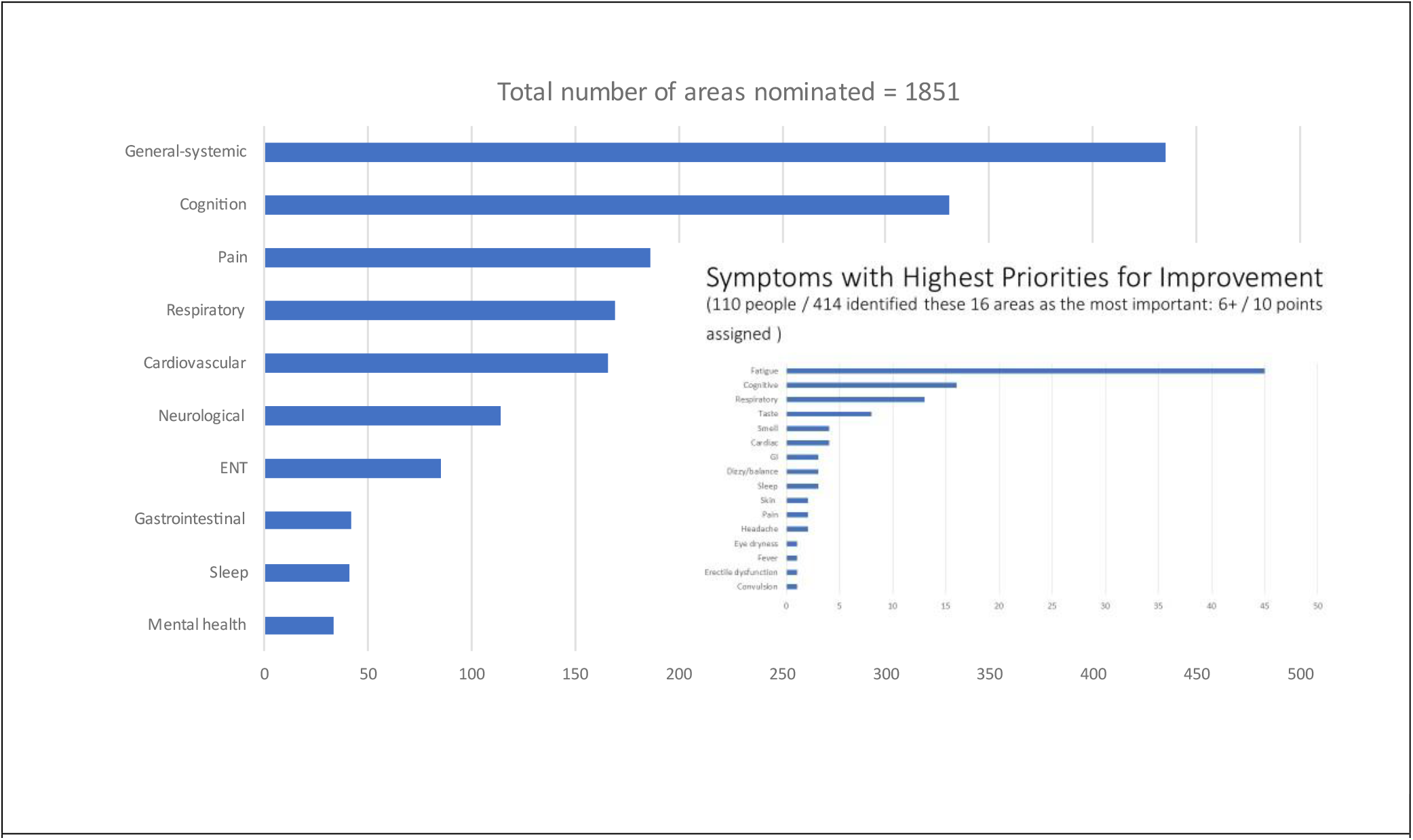
Most Frequently Nominated Symptoms according to Organ System and Symptoms with Highest Priority for Improvement

**Table 2** presents values on measures of symptoms. The fatigue construct was represented in four different ways. Fatigue severity was high, 65.3 / 100 with 100 as the worst fatigue. Over 80% reported symptoms indicative of PEM and over 1/3 identified that their fatigue required resting most of the day. One quarter of the sample reported symptoms indicative of PTSD and over 20% reported severe SOB affecting daily activities. Over 1/3 were prescribed medications for the health effects of PCS. Loneliness was reported by over 60% of the sample compared with 10.4% of the general Canadian population of similar age; 21.2% reported irritability.

**Table 2:** Symptom, Function, Health, and QOL Impact of PCS

| Symptom [Norm] | N or Mean [Median] | % or SD | Model |
| --- | --- | --- | --- |
| Fatigue |  |  |  |
| Severity* (VAS 0 -100 worst) | 65.3 [70] | 24.6 | Quantile |
| Post-exertional malaise (PEM)* | 347 (83.8%) |  | Logistic |
| Need to rest during the day |  |  |  |
| No | 38 | 9.2 | Referent |
| 1+ times during the day | 220 | 53.1 | Ordinal |
| Most of the day | 156 | 37.7 | Ordinal |
| Sleep* (VAS 0 -100 worst) | 50.0 [54.5] | 29.6 | Quantile |
| Distress |  |  |  |
| Severity (VAS 0 – 100 worst) | 38.8 [36.5] | 29.9 | Quantile |
| Men | 46.2 [50] | 31.5 |  |
| Women | 36.4 [30] | 29.1 |  |
| PTSD ($\geq 3/5$ symptoms) | 103 | 24.9 | Logistic |
| Shortness of breath (SOB) |  |  |  |
| Severity (VAS 0 – 100 worst) | 36.0 [30] | 29.2 | Quantile |
| Breathlessness |  |  |  |
| SOB walking 100 meters on level ground | 65 | 15.7% | Logistic |
| SOB dressing / can't leave house | 24 | 5.8% | Logistic |
| Prescribed medication for PCS symptoms | 153 | 37.0 | Logistic |
| Lonely sometimes or often [10.4%] | 254 | 61.4 | Logistic |
| Irritable often to always | 88 | 21.2 | Logistic |
| Cognition:C3Q (0-100 best) [81] | 43.5 [41.6] | 26.4 | Linear |
| Most prevalent symptoms of concentration loss |  |  |  |
| Too many thoughts in my head | 247 | 59.7 |  |
| Can't pay attention to two things at a time | 237 | 57.3 |  |
| Can't do complex tasks | 221 | 53.4 |  |
| Can't reading more than a few pages | 207 | 50.0 |  |
| Can't pay attention to verbal instructions | 188 | 45.4 |  |
| Most prevalent memory symptoms |  |  |  |
| Forgot what I was about to do | 245 | 59.2 |  |
| Forgot if what I have just read | 184 | 44.5 |  |
| Forgot tasks or activities I need to do | 180 | 43.5 |  |
| Forgot what I was supposed to buy at the store | 159 | 38.4 |  |
| Forgot what I have already done something | 157 | 37.9 |  |

Norms for C3Q are from Askari et al.(34)
Population rate of loneliness are from Statistics Canada (35)
\*age effect: older adults had lower symptom burden.

Cognitive function was poor with people reporting cognitive challenges in everyday life activities. On a self-report measure of cognitive concerns (C3Q), the average value out of 100 was 43.5 compared with 81 for a comparative sample. The most prevalent challenges were related to concentration with over 50% reporting challenges.

**Table 3** shows the impact of PCS on health and QOL outcomes. Values on the subscales (higher is better) of the RAND-36 were substantially lower than normative values as shown in Figure 4. There were sex effects for physical function, pain, social function, and role emotional subscales with women having lower values. Values on the PGI (mean: 26.3) are lower than for other methods of measuring QOL (means ≈38) as this format is based on people nominating only negative aspects of their life.

**Table 3.** Health-Related Quality of Life Outcomes

| HRQL Measures (0-100 best) | Norm (45-54 years) | Mean (SD) | Median [1QR] | Model |
| --- | --- | --- | --- | --- |
| **RAND-36** |  |  |  |  |
| Physical Function | 86.6 | 52.1 (27.9) | 50 [30-75] | Quantile |
| Men | 88.5 | 56.3 (29.7) | 65 [30-80] |  |
| Women | 89.3 | 51.0 (27.1) | 50 [30-70] |  |
| Role Physical (0 vs. >0) | 82.0 | 10.7 (24.9) | n=331/83 | Logistic |
| Bodily Pain | 72.9 | 45.8 (24.7) | 41 [31-62] | Quantile |
| Men | 77.5 | 50.1 (26.7) | 51 [31-72] |  |
| Women | 79.6 | 44.6 (23.7) | 41 [31-62] |  |
| General Health* | 77.2 | 41.6 (22.7) | 40 [25-57] | Linear |
| Vitality* | 63.3 | 23.1 (19.2) | 20 [10-35] | Quantile |
| Social Function media | 84.3 | 34.4 (26.4) | 25 [12.5-50] | Quantile |
| Men | 87.5 | 38.0 (27.2) | 37.5 [12.5-62.5] |  |
| Women | 88.5 | 33.6 (25.9) | 25.0 [12.5-50] |  |
| Role Emotional | 84.2 | 39.0 (43.3) | 33 [0-100] | Ordinal |
| Men (% ≤ median) |  |  | 71.3% |  |
| Women (% ≤ median) |  |  | 61.8% |  |
| Mental Health | 75.6 | 54.0 (20.9) | 56 [40-68] | Linear |
| **EQ-5D Index\*** (0 – 1 best) | 0.80 | 0.63 (0.23) | 0.69 [0.5 – 0.8] | Linear |
| **General health** |  | 38.5 (22.4) | 35.0 [21.0 – 50.0] | Linear |
| **QOL** |  | 37.9 (24.2) | 35.0 [20.0 – 50.0] | Linear |
| **PGI** |  | 26.3 (14.6) | 25.0 [17.0 – 34.0] | Linear |

\* age effect, older is better
Norms for RAND-36 are from Hopman et al.(36); Norms for EQ-5D Utility are from Poder et al.(37)

**Figure 2** shows the results of the cluster analysis. Five clusters were identified mainly distinguished by the degree of symptom burden. The effects of cluster membership on health and QOL outcome, estimated using linear regression with adjustment for age and sex, are shown in **Figure 3**. There was a significant cluster effect for all outcomes. Cluster membership explained over 30% of the variability in these health outcomes and there was an age effect only for the EQ-5D index value with older age associated with higher health utility.

**Figure 2.**
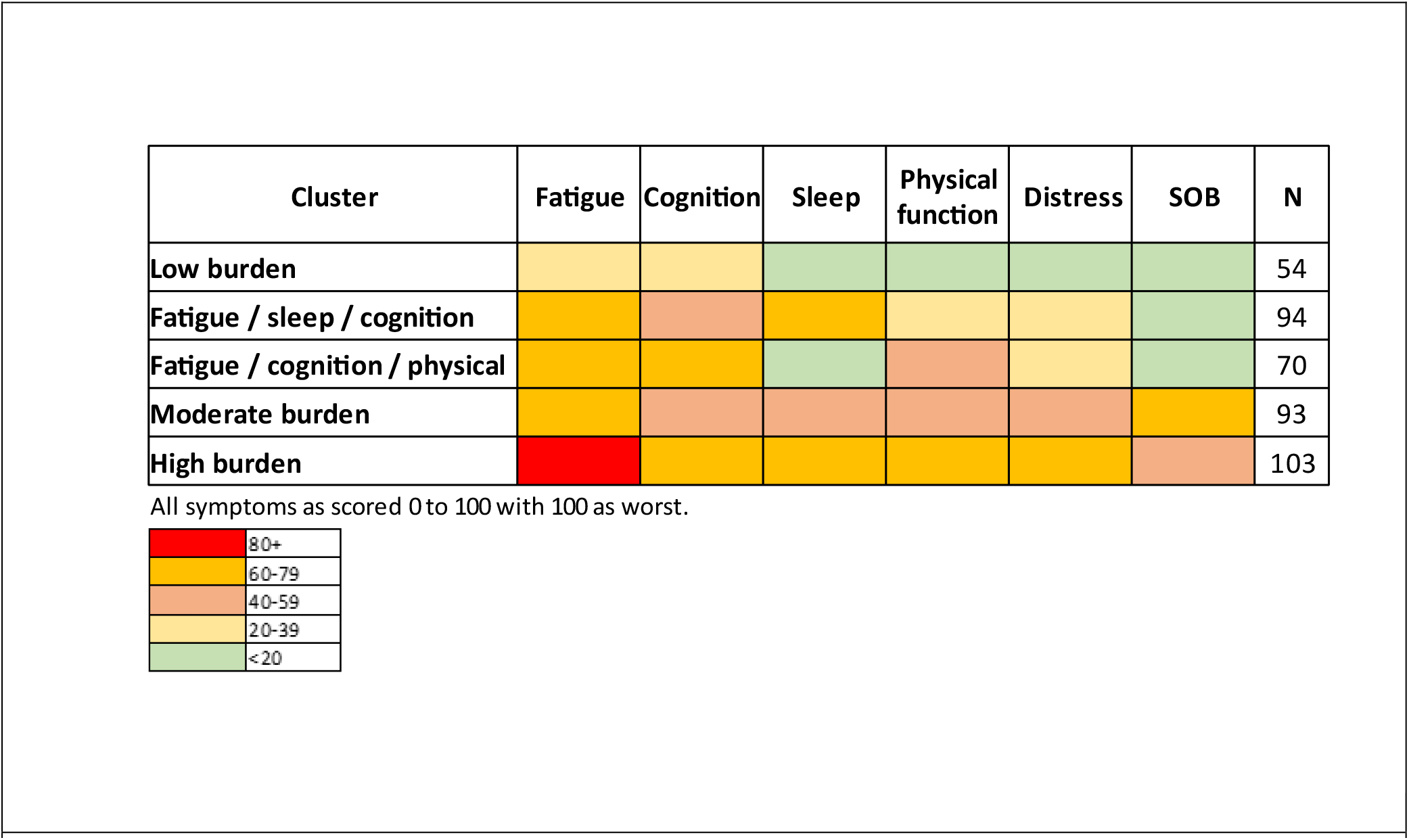
Symptom Clusters

**Figure 3.**
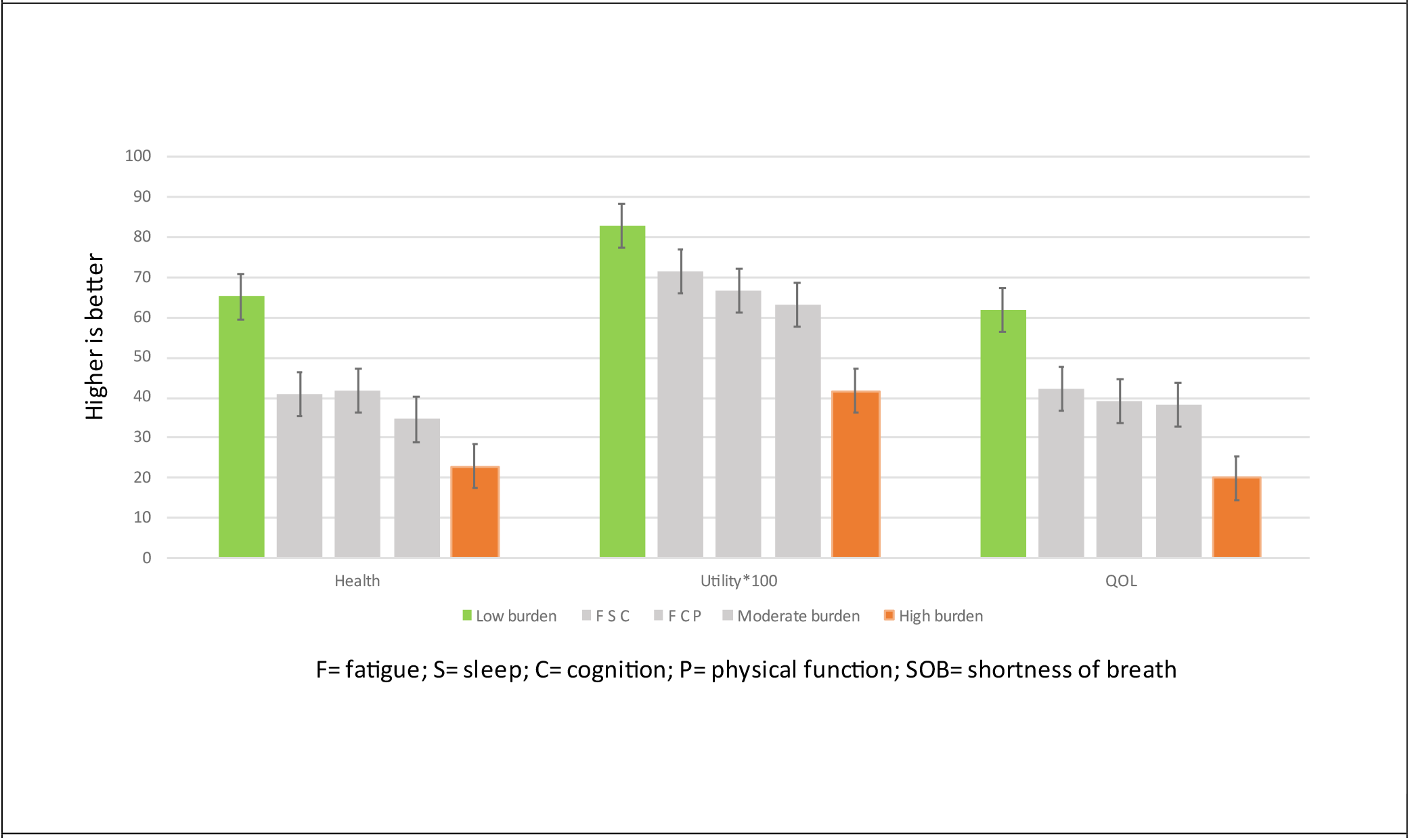
Impact of Symptom Cluster on Health and QOL Outcomes

**Figure 4.**
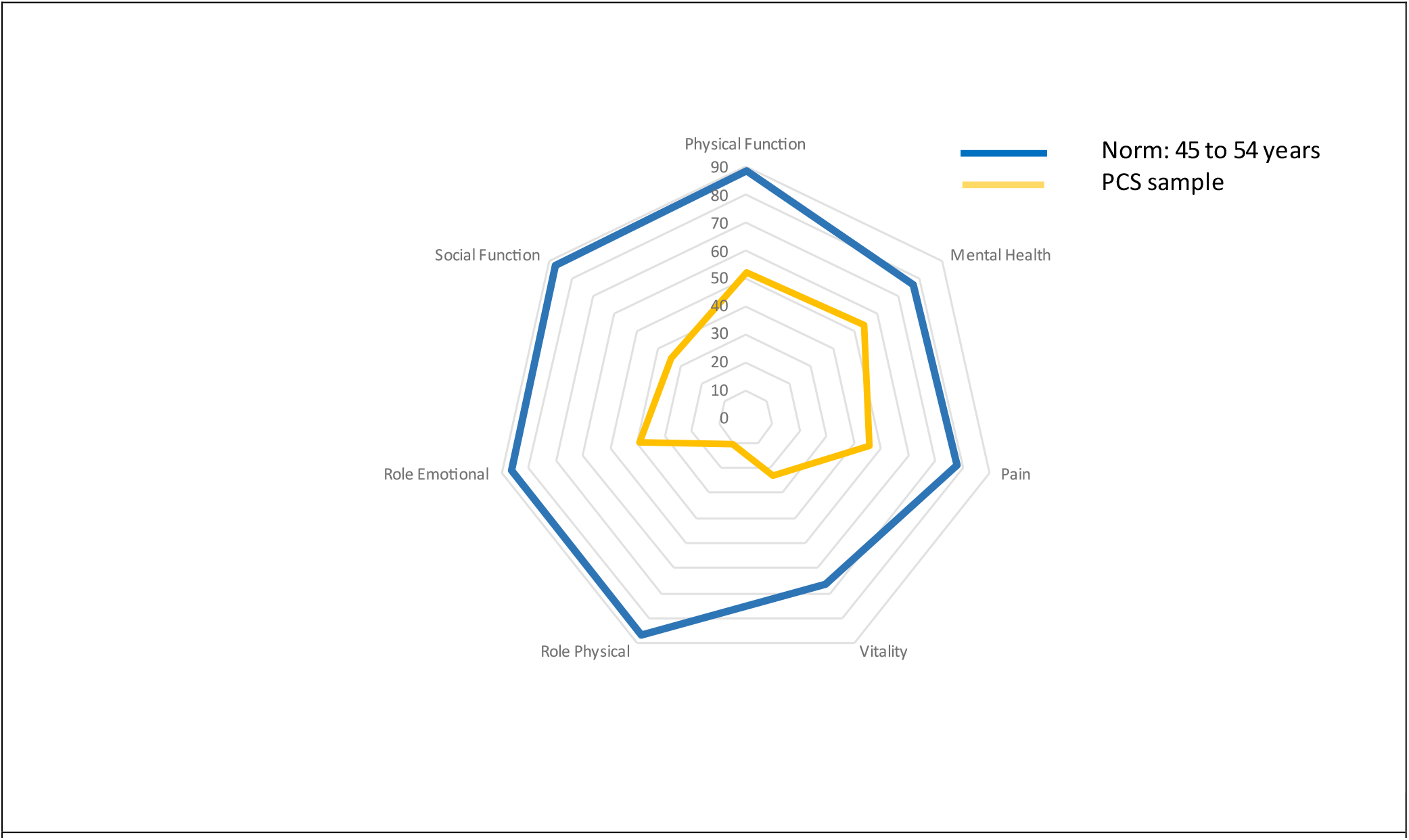
Values on RAND-36 Subscales for People with PCS (Yellow) and Normative Values (Blue) for people 45-54 years

### Interpretation

This study of adults self-identifying with PCS in Quebec, Canada found long-lasting, widespread, and severe effects across the full range of health outcomes (symptoms, function, health perception, and QOL). The symptom of most concern to this sample was fatigue, with over 80% reporting PEM and over 90% needing to rest during the day. Cognitive symptoms were also very prevalent with people scoring 40 (out of 100) points lower than expected on a measure of self-reported cognitive concerns.

Concentration difficulties were the most common cognitive issue. Respiratory symptoms were also frequent. Symptoms had substantial functional impact, with close to half the sample on sick leave. Health-related quality of life as assessed with the EQ-5D index values for the QAPC sample was 0.63, comparable to values reported for Canadians with stroke, multiple sclerosis, or advanced cancer(17, 18) and far below the normative value for Quebec (0.80).(37)

A similar profile of PCS burden has been reported by others(15, 23, 40-45) although it is difficult to compare studies owing to the differences in how samples were assembled. PCS is by definition a self-reported syndrome and hence how people are recruited into studies will affect estimates of symptom burden. The QAPC sample was predominately women (76%), as in other studies with self-identified PCS samples(46). In contrast, in the UK census study that is the best available population-level data on self-reported PCS to date (6), the women to men ratio was ≈3:2. However, we found that women and men were similar on the outcomes we assessed. We observed effects of age on some variables.

Paradoxically, while older age was associated with higher likelihood of hospitalization during acute infection, it was associated with lower PCS symptom burden and better QOL.

We assessed symptom clustering, seeking evidence for sub-syndromes that might have different etiologies or contributors. The observed pattern is consistent with a range of severity rather than sub-syndromes, with fatigue an early feature in even the mildest stages, and additional symptoms accruing with increasing severity. A recent systematic review of 151 studies from 32 countries with a total of 1,285,407 participants who would meet criteria for PCS(12) reported that the proportion of people with at least one symptom decreased over time from 56.0% (at 1–3 months) to 37.8% (at 6–12 months). This indicates that over a third of people reporting PCS can be symptomatic for a year or more. While the cross-sectional observations here do not address the evolution of PCS, participants had symptoms for nearly a year at study entry, on average. On-going longitudinal follow-up will shed light on the evolution and day-to-day variability in this seemingly chronic phase of COVID-19.

This study is affected by the same limitations as much of the existing literature on PCS, as there is no base population to sample from and people entering the study were likely those with the most severe and persistent symptoms. Further, while over 90% of the QAPC sample reported a positive COVID test, infection status was not independently verified. There was widespread availability of PCR testing in the early waves of the pandemic in Quebec, and home tests were provided free of charge in pharmacies thereafter. Study participants likely had SARS-CoV-2 infection with persistent disabling chronic symptoms and most likely to seek help.

While the findings from this self-identified sample do not provide information on prevalence, they can inform healthcare service planning: People identified relief from fatigue, pain, and from cognitive, respiratory, and cardiac symptoms as their top priorities for treatment. This information could help prioritize current management recommendations which are based on reported frequency and expert opinion.(47) Rehabilitation approaches may be particularly helpful, given the nature, functional impact, and chronicity of the most bothersome symptoms. Most people had multiple symptoms, indicating that a multidisciplinary approach to assessment and treatment is warranted. Management of the most common chronic symptoms falls within the scope of practice of physical therapy, occupational therapy, psychology, and nursing. Social services addressing the financial and functional impact of PCS will also be important.

These initial results from the QAPC study provide good information as to the range of problems experienced, their severity, and patient-centered priorities for intervention in the Canadian context. There is a need for people with PCS to have access to clinical care for management of complications, rehabilitation, as well as support for self-management of symptoms if appropriate. Further work is needed to identify the most effective interventions and to identify risk and resilience factors in the evolution of symptoms over time.

## Data Availability

All data produced in the present study will be available online shortly

